# International Validation of Echocardiographic AI Amyloid Detection Algorithm

**DOI:** 10.1101/2024.12.14.24319049

**Authors:** Grant Duffy, Evangelos K. Oikonomou, Jonathan Hourmozdi, Hiroki Usuku, Jigesh Patel, Yoshinori Katsumata, Wakako Yamasawa, Lily Stern, Shinichi Goto, Kenichi Tsujita, Rohan Khera, Faraz S. Ahmad, David Ouyang

**Author notes:** **Correspondence:** David Ouyang, MD, 127 S. San Vicente Blvd., Suite A3600, Los Angeles, CA 90048.

## Abstract

**Background:** Diagnosis of cardiac amyloidosis (CA) is often missed or delayed due to confusion with other causes of increased left ventricular wall thickness. Conventional transthoracic echocardiographic measurements like global longitudinal strain (GLS) have shown promise in distinguishing CA, but with limited specificity. We conducted a multi-site retrospective case-control study to investigate the performance of a computer vision algorithm for CA identification across multiple international sites.

**Methods:** EchoNet-LVH is a computer vision deep learning algorithm for the detection of cardiac amyloidosis based on parasternal long axis and apical-4-chamber view videos. We evaluated EchoNet-LVH’s ability to distinguish between the echocardiogram studies of CA patients and controls. We reported discrimination performance with an area under the receiver operating characteristic curve (AUC) and associated sensitivity, specificity, and positive predictive value at the pre-specified threshold.

**Results:** EchoNet-LVH had an AUC of 0.896 (95% CI 0.875 – 0.916). At the pre-specified model threshold optimizing for specificity, EchoNet-LVH had a sensitivity of 0.644 (95% CI 0.601 – 0.685), specificity of 0.988 (0.978 – 0.994), positive predictive value of 0.968 (95% CI 0.944 – 0.984), and negative predictive value of 0.828 (95% CI 0.804 – 0.850). There was no evidence of heterogeneity in performance by site, race, sex, age, BMI, CA subtype, or ultrasound manufacturer.

**Conclusion:** EchoNet-LVH can assist with earlier and accurate diagnosis of CA. EchoNet-LVH achieved development goals to be highly specific to maximize positive predictive value of downstream confirmatory testing since CA is a rare disease.

## INTRODUCTION

Cardiac amyloidosis (CA) is caused by deposition of misfolded proteins in the myocardium, including transthyretin (ATTR) or immunoglobulin light chains (AL)^1,2^. Regardless of the etiology, CA leads to increased left ventricular wall thickness and heart failure, however early symptoms can be non-specific and not readily recognized^3–5^. Common echocardiographic measurements are insufficient to precisely discriminate CA from other etiologies of left ventricular hypertrophy (LVH) or heart failure^6–8^.

There is concern that CA is underdiagnosed and diagnosed too late, which limit the opportunity to receive recent targeted therapies that improve quality of life and decrease mortality outcomes in CA patients^9–11^. Recent research has focused on methods that can assist with early identification of CA^12–14^. Echocardiography is one of the most common initial tests when evaluating patients with heart failure symptoms, with typical CA features on including increased left ventricular wall thickness, normal or small left ventricular cavity, preserved left ventricular ejection fraction (LVEF), and diastolic dysfunction^1,2^. However, many of these features are also commonly found in other forms of heart failure with preserved ejection fraction and have limited specificity to identify CA^6,8^, resulting hesitancy to highlight suspicion for CA.

Recent advances in computer vision and artificial intelligence (AI) have enabled precision phenotyping of structure and function in cardiac ultrasound as well as other forms of cardiovascular diagnostics^15–19^. AI applied to echocardiography can precisely estimate wall thickness^13^, assess mitral regurgitation severity^20^, and left ventricular ejection fraction (LVEF)^21^ as well as detect cardiac amyloidosis^13^, HCM, and diastolic dysfunction^22^. EchoNet-LVH is one such computer vision model developed for the detection of CA^13^. An automated pipeline, EchoNet-LVH identifies parasternal long axis and apical-4-chamber views from transthoracic echocardiogram studies to precisely measure wall thickness and identify texture and motion suggestive of CA. Trained against other forms of left ventricular hypertrophy as controls against CA cases, EchoNet-LVH was designed to be specific in discriminating between CA and phenotypic mimics.

In this study, we evaluated the performance EchoNet-LVH across multiple new healthcare systems on videos that the model has never seen before. We conducted a multi-site international retrospective case-control study evaluating EchoNet-LVH’s ability to distinguish between the echocardiogram studies of CA patients and controls. We reported discrimination performance with area under the receiver operating characteristic curve (AUC) and associated sensitivity, specificity, and positive predictive value at a pre-specified threshold to identify CA.

## METHODS

### Study Design

We conducted an international multicenter retrospective case-control cohort study with participants from multiple geographically distinct healthcare systems to evaluate EchoNet-LVH. EchoNet-LVH was previously developed using CA cases and controls from Stanford Healthcare^20^, so this study serves as temporally and geographically distinct external validation. Patients were sourced from Cedars-Sinai Medical Center in Los Angeles, California, Keio University in Tokyo, Japan, Northwestern Medicine in Chicago, Illinois, and Yale-New Haven Hospital in New Haven, Connecticut. A total of 520 patients were identified as having CA and matched to 903 randomly selected controls from patients receiving echocardiography and at least 65 years of age. CA patients were diagnosed with transthyretin (ATTR), light chain (AL) amyloidosis, or other forms of cardiac amyloidosis using a combination of nuclear cardiac amyloid imaging (including SPECT/CT with approved bone radiotracers, such as Tc^99m^-pyrophosphate), monoclonal gammopathy testing, genetic testing, and/or tissue biopsy. The controls either had negative testing for cardiac amyloidosis or testing was not performed. Given the low population prevalence of CA, the likelihood of undiagnosed CA in the controls was thought to be minor and unlikely to change the analysis. This study was approved by the Cedars-Sinai Institutional Review Board.

### Computer Vision Model

EchoNet-LVH’s development approach and internal validation has been previously described^20^. In short, EchoNet-LVH is an automated machine learning pipeline that automatically identifies parasternal long axis and apical-4-chamber views from transthoracic echocardiogram studies, precisely measures wall thickness, and assesses texture and motion from the apical-4-chamber view echocardiogram videos to assess suspicion for cardiac amyloidosis. Information from the apical-4-chamber view is synthesized with a segmentation model’s assessment of wall thickness from the parasternal long axis videos to come up with a suspicion for CA. Given the low population prevalence of CA, a prespecified threshold (0.8) in the summative assessment was chosen to optimize and prioritize for specificity, which in turn, maximizes positive predictive value. Echocardiogram videos were obtained in DICOM format and a fully automatic pipeline analyzed the study. All sites ran the model independently without transfer of imaging data through the sharing of a Docker container with code for the full end-to-end pipeline.

### Statistical Analysis

Continuous variables were reported using median (interquartile range), and categorical variables were reported with number (percentage). Performance of in the model for discriminating CA was evaluated using area under the receiver operator curve (AUC), sensitivity, specificity, positive predictive value (PPV), and negative predictive value (NPV). Ninety-five percent confidence intervals were assessed for all analyses. Statistical analysis was performed in Python (Python Software Foundation, Beaverton, Oregon).

## RESULTS

Demographics and clinical characteristics of the study cohort are shown in **Table 1**. The mean age of the cohort was 78.2 (IQR 72 – 84) and 77.7% male, with cases matched 2:1 with controls greater than 65 years of age. In the cases, there was representation from both AL (22.7%) and ATTR (74.2%) amyloidosis. There was a wide range of age, BMI, and ultrasound manufacturers across the sites with subgroup analysis to evaluate for heterogeneity of EchoNet-LVH performance..

**FIGURE 1.**
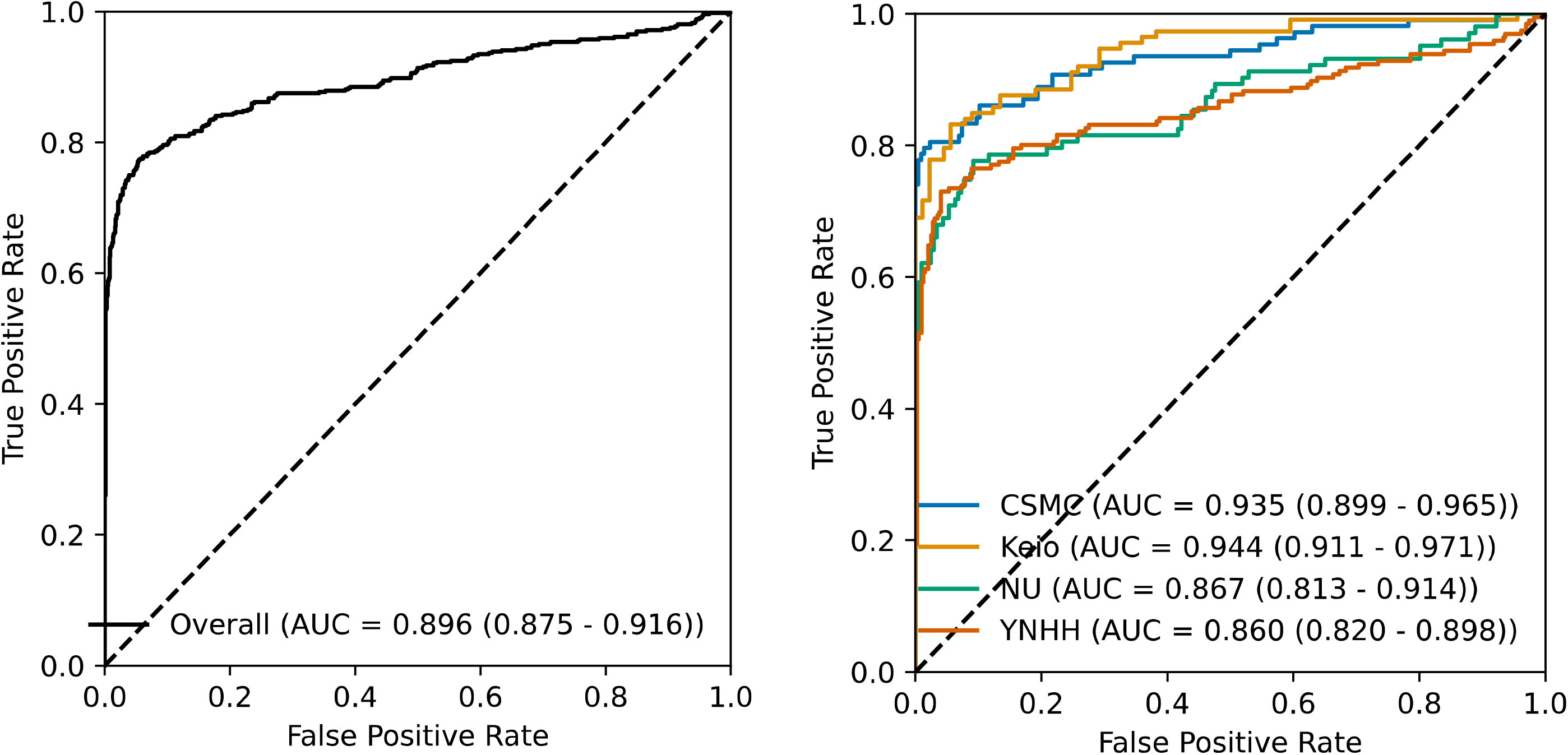
Receiver Operating Characteristic Curve for EchoNet-LVH. Overall performance of AI algorithm and subset by site. CSMC = Cedars-Sinai Medical Center, NU = Northwestern University, YNHH = Yale New Haven Hospital

**Table 1.** Patient Characteristics.

| Participants | Median (IQR) or N (%) |
| --- | --- |
| Age | 78 (72-84) |
| Men | 1105 (77.7%) |
| Cardiac amyloidosis | 231 (33.3%) |
| Subtype |  |
| AL | 118 (22.7%) |
| ATTR | 386 (74.2%) |
| Other | 16 (3.2%) |
| Age |  |
| < 74 | 456 (32.0%) |
| 75-84 | 626 (44.0%) |
| > 85 | 341(24.0%) |
| BMI |  |
| < 25 | 604 (42.4%) |
| 25-30 | 511 (35.9%) |
| > 30 | 308 (21.6%) |
| Ultrasound Manufacturer |  |
| Philips | 973 (68.4%) |
| GE | 281 (19.7%) |
| Siemens | 130 (9.1%) |
| Toshiba | 39 (2.7%) |

EchoNet-LVH had an overall AUC of 0.896 (95% CI 0.875 - 0.916) with minimal site level variation in performance (**Table 2**). The lowest site AUC was YNHHS (Yale-New Haven Health System) with an AUC 0.860 (95% CI 0.818 – 0.898) and the highest site AUC was Keio University with an AUC of 0.944 (95% CI 0.911 – 0.971). The overall sensitivity was 0.644 (95% CI 0.601 – 0.685) and the overall specificity was 0.988 (95% CI 0.978 – 0.994). There was no significant heterogeneity in other performance characteristics across site. At a 2:1 ratio of controls to cases, EchoNet-LVH had a PPV of 0.968 (95% CI 0.944 – 0.984) and a NPV of 0.828 (95% CI 0.804 – 0.850).

**Table 2.** Overall performance. CSMC = Cedars-Sinai Medical Center, NU = Northwestern University, YNHH = Yale New Haven Hospital, PPV = Positive Predictive Value, NPV = Negative Predictive Value

| Group | n | Sensitivity (95% CI) | Specificity (95% CI) | PPV (95% CI) | NPV (95% CI) | AUROC (95% CI) |
| --- | --- | --- | --- | --- | --- | --- |
| Overall | 1423 | 0.644 (0.601 - 0.685) | 0.988 (0.978 - 0.994) | 0.968 (0.944 - 0.984) | 0.828 (0.804 - 0.850) | 0.896 (0.875 - 0.916) |
| CSMC | 324 | 0.648 (0.550 - 0.738) | 1.000 (0.983 - 1.000) | 1.000 (0.949 - 1.000) | 0.850 (0.800 - 0.892) | 0.935 (0.899 - 0.965) |
| Keio | 202 | 0.779 (0.691 - 0.851) | 0.966 (0.905 - 0.993) | 0.967 (0.907 - 0.993) | 0.775 (0.686 - 0.849) | 0.944 (0.911 - 0.971) |
| NU | 309 | 0.621 (0.520 - 0.715) | 0.981 (0.951 - 0.995) | 0.941 (0.856 - 0.984) | 0.838 (0.785 - 0.882) | 0.867 (0.814 - 0.915) |
| YNHH | 588 | 0.577 (0.504 - 0.647) | 0.990 (0.974 - 0.997) | 0.966 (0.915 - 0.991) | 0.824 (0.786 - 0.857) | 0.860 (0.818 - 0.898) |

EchoNet-LVH also had similar performance across patient characteristics and ultrasound manufacturer (**Table 3**). EchoNet-LVH had an AUC of 0.921 (95% CI 0.882 - 0.955) for detecting AL cardiac amyloidosis and an AUC of 0.887 (95% CI 0.862 - 0.911) for detecting ATTR cardiac amyloidosis. Our model had similar performance for men (AUC of 0.893 [95% CI 0.869 – 0.914]) and women (AUC of 0.904 [95% CI 0.849 – 0.950]). There was no significant heterogeneity by race, age, BMI, or ultrasound manufacturer. Across all key groups, there was similar sensitivity and specificity of our approach, and there was no trend for differences in performance across subclasses.

**Table 3.**
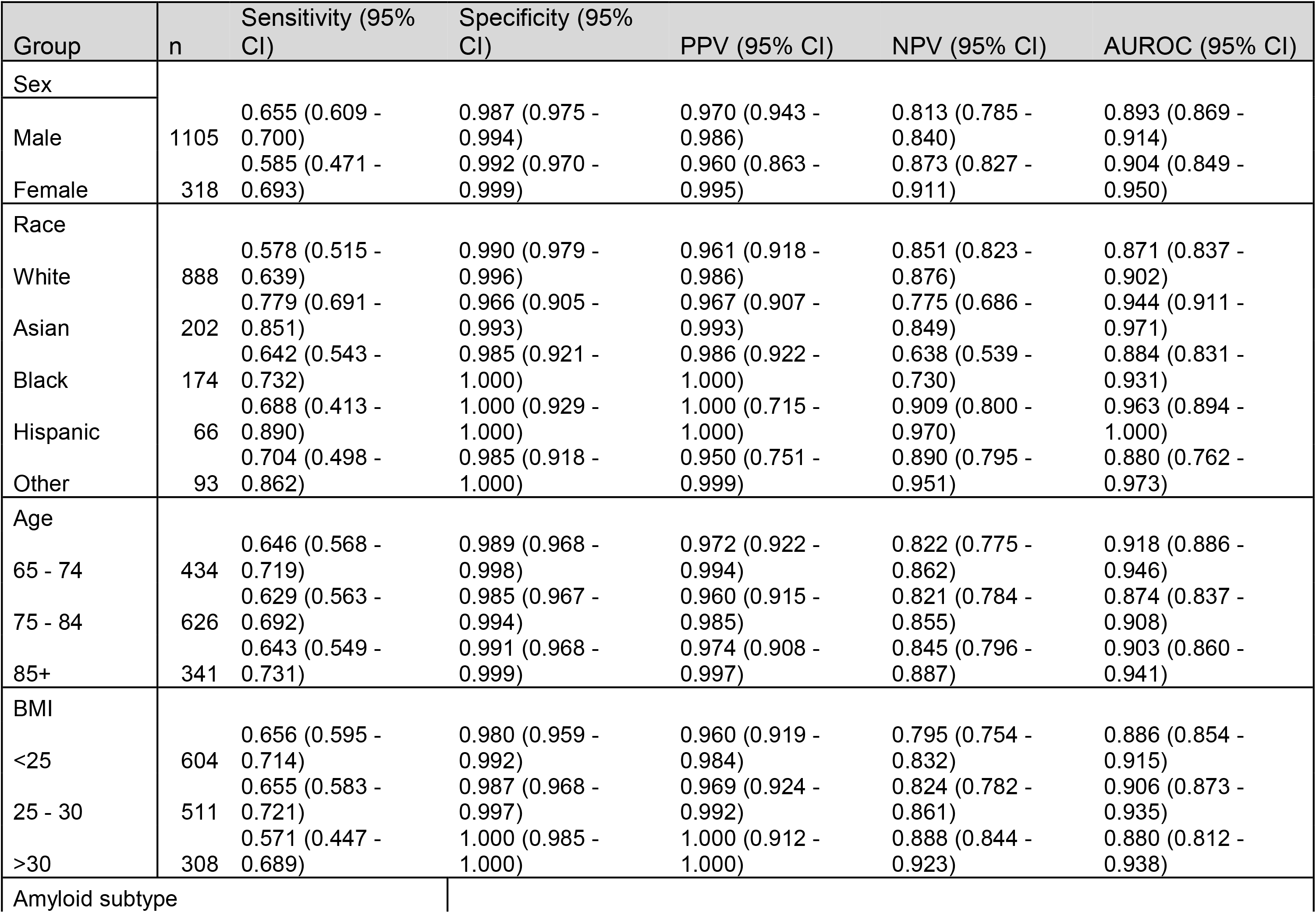
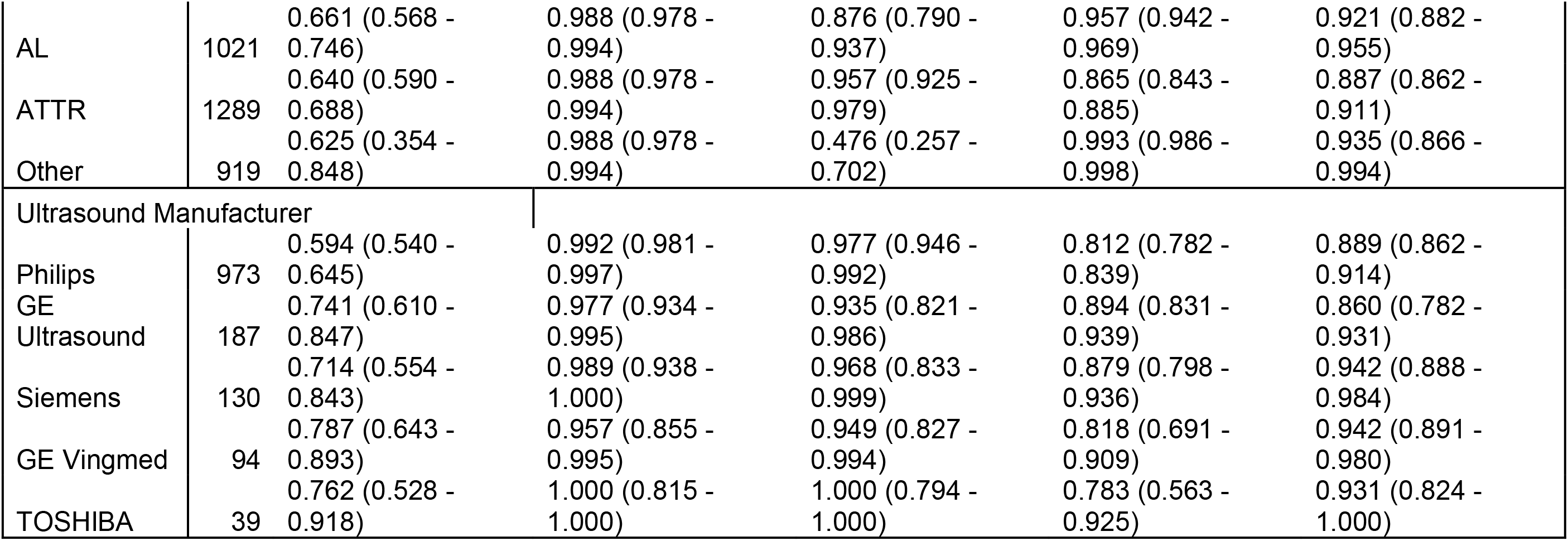
Subgroup performance. PPV = Positive Predictive Value, NPV = Negative Predictive Value, BMI = Body Mass Index

## DISCUSSION

In this study, we evaluated the performance of a computer vision driven AI workflow for the detection and screening of cardiac amyloidosis across a wide range of patients from an international cohort of geographically distinct healthcare systems. In this setting, we found EchoNet-LVH had strong performance in identifying patients with CA of all subtypes, and its performance was consistent across sites, ultrasound manufacturers, and patient characteristics.

A few things are worth considering in evaluating our algorithm. Because increased wall thickness is a hallmark of CA^1,23^, our algorithm automated the approach to precisely measuring wall thickness as well as integrated this precise measurement with more ‘black box’ features of motion and texture assessed in the apical-4-chamber view. Our approach homes to minimize or exclude confounders^13^, as our models were trained on controls matched on wall-thickness and limit the potential of AI models to shortcut on wall thickness alone. Second, our approach sought to maximize positive predictive value in the downstream testing of CA. In a rare disease, PPV is significantly impacted by model specificity (as the number of false positives are likely to outweigh the number of potential true positives). In the algorithm development, we optimized for specificity as to minimize the number of false positives rejected in downstream testing.

A few limitations are worth considering as further study is still warranted. Prospective trialing of CA screening approaches has not yet completed. There is incomplete data on the true population prevalence of CA^3–5,24^, which significantly impacts the PPV of any algorithm. Multiple other measurements and approaches have been suggested to screen for CA^25–27^, however our results represent one of the few fully automated pipelines. Additionally, other work has assessed the performance of EchoNet-LVH in comparison to score-based decision aids and EHR-based algorithms and show EchoNet-LVH’s superior performance^27^. Furthermore, there is significant observer variability in most echocardiographic measurements^28^, which limit the precision of approaches based on routine echo measurements alone^8^.

In this study comparing CA patients with patients who were referred for pyrophosphate scintigraphy but had CA ruled out, we demonstrate that IVSd/GLS can be used to identify CA in a population with high clinical suspicion for CA. This ratio has superior performance compared to individual echocardiographic measurements as well as other ratios that are already abnormal in patients at high suspicion for CA. If validated in future studies, incorporation of this easily obtainable measurement can assist with earlier diagnosis of CA resulting in reduced morbidity and mortality.

## Data Availability

All data produced in the present study are available upon reasonable request to the authors

## Disclosures

This work is funded by NIH NHLBI grants R00HL157421, R01HL173526, and R01HL173487 to DO, and a grant from Alexion AstraZeneca Rare Disease. GD is currently an employee of Meta. D.O. reports consulting fees and/or equity in Ultromics, InVision, EchoIQ, and Pfizer. R.K. reports support from NIH NHLBI (R01HL167858 and K23HL153775), NIH NIA (R01AG089981), and the Doris Duke Charitable Foundation (2022060). He is an Associate Editor of JAMA and receives research support, through Yale, from the Blavatnik Foundation, Bristol-Myers Squibb, Novo Nordisk, and BridgeBio. He is a coinventor of Pending Patent Applications WO2023230345A1, US20220336048A1, 63/346,610, 63/484,426, 63/508,315, 63/580,137, 63/606,203, 63/619,241, and 63/562,335, and a co-founder of Ensight-AI, Inc and Evidence2Health, LLC. E.K.O. is supported through NIH NHLBI (F32HL170592). He is a co-founder of Evidence2Health LLC, an ad hoc consultant for Caristo Diagnostics Ltd and Ensight-AI Inc, and has received royalty fees from technology licensed through the University of Oxford, outside this work. FSA has received research support from Pfizer and Atman Health. All other authors declare no competing interests.

## Notes

### Author Declarations

IRB of Cedars Sinai Medical Center gave ethical approval for this work

### Summary of Updates

Add authors involved in external validation

